# Stereotactic arrhythmia radioablation for refractory scar-related ventricular tachycardia

**DOI:** 10.1101/19012880

**Authors:** Carola Gianni, Douglas Rivera, J David Burkhardt, Brad Pollard, Edward Gardner, Patrick Maguire, Paul C Zei, Andrea Natale, Amin Al-Ahmad

## Abstract

**Background:** Stereotactic radiosurgery is a form of radiotherapy that is performed in a single session and focuses high-dose ionizing radiation beams from a collimated radiation source to a small, localized area of the body. Recently, stereotactic radiosurgery has been applied to arrhythmias (stereotactic arrhythmia radioablation - STAR), with promising results reported in patients with refractory, scar-related ventricular tachycardia (VT), a cohort with known high morbidity and mortality.

**Objective:** Herein, we describe our experience with the use of CyberKnife, a frameless image-guided linear accelerator stereotactic radiosurgery system, in conjunction with CardioPlan, a cardiac specific radiotherapy planning software, to treat patients with scar-related VT, detailing its early and mid-to long-term results.

**Methods:** This is a pilot, prospective study of patients undergoing STAR for refractory VT. The anatomical target for radioablation was defined based on the clinical VT morphology, electroanatomical mapping, and study-specific pre-procedural imaging with cardiac computed tomography. The target volume delineated with the aid of CardioPlan was treated with a prescription radiation dose of 25 Gy delivered in a single fraction by CyberKnife in an outpatient setting. Ventricular arrhythmias and radiation-related adverse events were monitored at follow-up to determine STAR efficacy and safety.

**Results:** Five patients (100 % male, 63 ± 12 years old, 80 % ischemic cardiomyopathy, left ventricular ejection fraction 34 ± 15 %) with refractory VT underwent STAR between January and June 2018. Radioablation was delivered in 82 ± 11 minutes without acute complications. During a mean follow-up of 12 ± 2 months, all patients experienced clinically significant mid-to late-term ventricular arrhythmia recurrence; two patients died of complications associated with their advanced heart failure. There were no clinical or imaging evidence of radiation necrosis or other radiation-induced complications in the organs at risk surrounding the scar targeted by radioablation.

**Conclusion:** Despite good initial results, STAR did not result in effective ventricular arrhythmia control in the long term in a selected, high-risk population of patients with scar-related VT. The safety profile was confirmed to be favorable, with no radiation-related complications observed during follow-up. Further studies are needed to explain these disappointing results.

## Introduction

Stereotactic radiosurgery is a form of radiotherapy that is performed in a single session and focuses high-dose ionizing radiation beams from a collimated radiation source to a small, localized area of the body. In contrast to traditional radiotherapy which functions by disrupting cellular division, radiosurgery works by creating radiation-induced necrosis.^1^ Stereotactic radiosurgery is currently used for treatment of various types of tumors in the head and neck, lung, abdomen, pelvis and spine, with good local control rates and minimal side effects.^2^ Stereotactic arrhythmia radioablation (STAR) is the application of stereotactic radiosurgery for the non-invasive treatment of cardiac arrhythmias. STAR has been used to homogenize the scar in patients with structural heart disease and ventricular tachycardia (VT) that is refractory to antiarrhythmic drugs (AADs) and has failed radiofrequency catheter ablation (RFCA), with promising results.^3-10^ In this report, we describe our experience with the use of CyberKnife, a frameless image-guided linear accelerator stereotactic radiosurgery system, in conjunction with CardioPlan, a cardiac specific radiotherapy planning software, to treat patients with scar-related VT, detailing its early and mid-to long-term results.

## Methods

### Patient population

This was a prospective, single-arm, two-center feasibility study of STAR for the treatment of refractory scar-related VT. Patients referred by an electrophysiologist to our center to be considered for radioablation were screened and considered for the study if they met all the following criteria: i) presence of an implantable cardioverter-defibrillator (ICD); ii) ischemic or non-ischemic cardiomyopathy with recurrent symptomatic VT that induced ICD shock(s) despite RFCA and/or AAD therapy; iii) age ≥ 60 years; iv) left ventricular ejection fraction (LVEF) ≥ 20%. Patients were excluded in case of: i) prior radiation therapy to the chest; ii) active ischemia or other reversible causes of VT; iii) disease process(es) likely to limit survival to < 12 months; iv) baseline creatinine levels > 1.5 mg/dL.

In this study, we report the clinical outcomes of patients enrolled in the CardioPlan software feasibility study, sponsored by CyberHeart (NCT02661048). All patients provided written informed consent and the study was approved by each participating center’s Institutional Review Board.

### Radioablation treatment plan

The anatomical target for radioablation was defined based on study-specific, pre-procedural imaging with cardiac computed tomography (CT). The latter consisted of an end-expiration, chin to navel chest CT scan, with a resolution of 1 to 1.25 mm axial slices, and contrast with maximal enhancement at the level of the pulmonary veins. CT imaging was performed after placement of a transjugular temporary active fixation pacing lead (Oscor PY58PV in the first three patients, Medtronic 5076-58 in the last two) in the right ventricular septal area, which served as the fiducial marker to dynamically track respiratory motion during delivery of radiotherapy (see below). To confirm that the area of myocardial thinning identified on contrast enhanced first-pass CT was the clinical arrhythmogenic target, it was correlated with low voltage areas seen on electroanatomical mapping performed during previous RFCA along with the site of origin (exit) of the clinical VT morphology on the 12-lead ECG.

The 3-dimensional contours of the myocardial substrate to be targeted by radioablation were at first created by the electrophysiologist with the CardioPlan software (CyberHeart, Portola Valley, CA, USA; Figure 1). This target, also referred to as the clinical target volume (CTV), included the whole transmural myocardium encompassing the scar. A margin of around 3 mm was added to the CTV to account for its motion relative to the cardiac cycle, thus defining the internal target volume (ITV). In addition, a contour of surrounding normal organs (organs at risk, OAR) was also delineated to define their relationship with the CTV. OAR taken into consideration for ventricular STAR included the mitral valve, coronary arteries outside of the scar, lung, large bronchi, esophagus, great vessels, stomach, liver, bowels, skin, and spinal cord. The ITV and OAR delineated with CardioPlan as well as raw CT images were then transferred to the MultiPlan Treatment Planning Software for Cyberknife (Accuray, Sunnyvale, CA, USA), in which the final target volume (or, planning target volume - PTV) was refined by the radiation oncologist, in collaboration with the electrophysiologist and nuclear medicine, to account for set-up uncertainties (Figure 1). For OARs, pre-defined radiation dose and volume limits were applied to minimize the probability of radiation-related toxicity, following Radiation Therapy Oncology Group (RTOG) recommendations based on published reports and radiobiological models for Normal Tissue Complication Probability (NTCP).^11^

**Figure 1.**
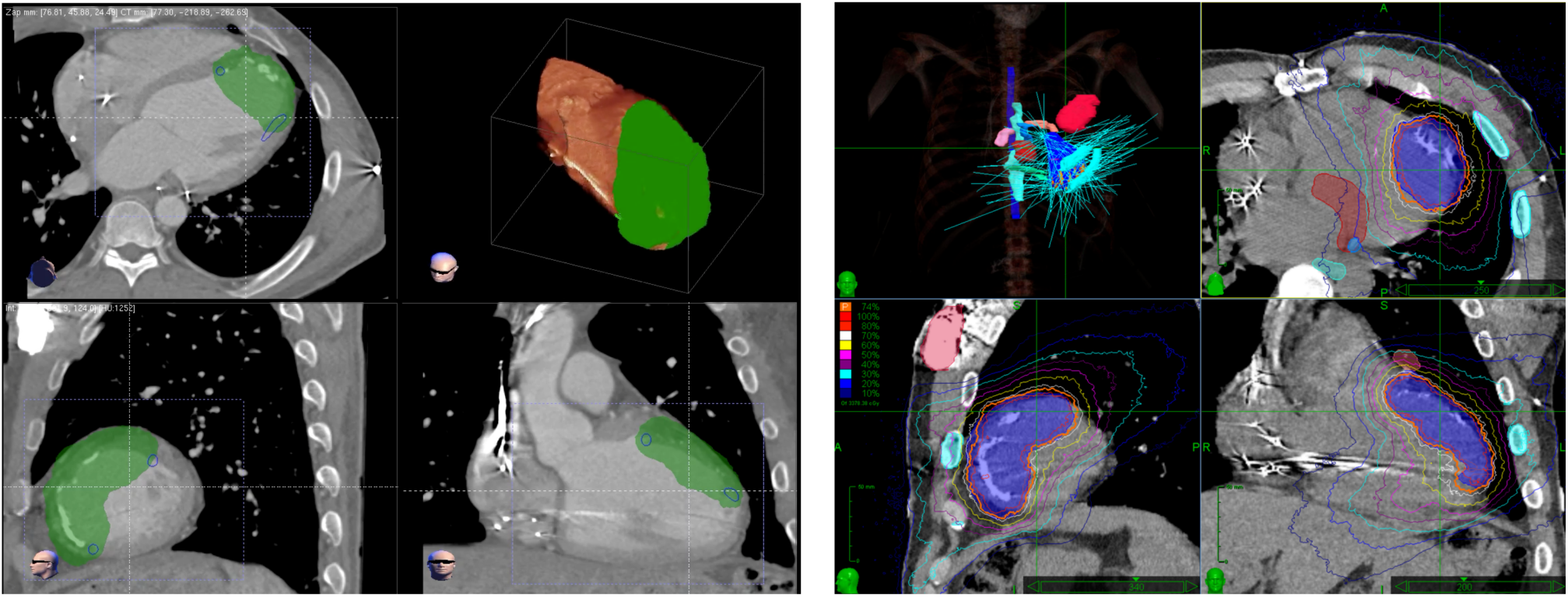
Treatment plan for radioablation of a left ventricular myocardial scar. Left, contouring of the CTV (green) created by the electrophysiologist with CardioPlan. Right, isodose distribution with PTV (blue) and its prescription isodose line (orange) created with MultiPlan Treatment Planning Software. Surrounding isodose lines are color coded according to the percentage of the prescription dose (legend on mid-left), thus illustrating the dose fall off outside the PTV. Selected adjacent OARs are also highlighted, including mitral valve (red), coronary arteries (pink, azure, orange), esophagus and ribs (turquoise), spinal cord (blue). CTV, clinical target volume; OAR, organ at risk; PTV, planning target volume

### Radioablation procedure

The final treatment plan was then transferred to a 6-MV linear accelerator (CyberKnife G4 system with an Iris variable aperture collimator) and the patient treated with a prescription dose of 25 Gy delivered in a single fraction to the PTV. CyberKnife is a frameless, image-guided radiotherapy delivery system in which the linear accelerator is mounted on a multijointed robotic arm able of aiming highly focused 6-MV radiation beams with six degrees of freedom (Figure 2).^12^ To accurately deliver radiotherapy, CyberKnife is equipped with a set of orthogonal X-ray sources, which acquire periodical images throughout treatment duration to monitor the position of the patient and locate the target within, adjusting for movements. For STAR, radiation was delivered using two motion tracking systems: X-Sight Spine Tracking System and Synchrony Respiratory Tracking. The former enables tracking of skeletal structures of the spine for accurate patient positioning, accounting for static rotational alignment.^13^ The latter creates a real-time model that correlates the motion of the fiducial marker placed near the PTV to the motion of the chest wall, accounting for dynamic (respiration-related) translational aligment.^14^

**Figure 2.**
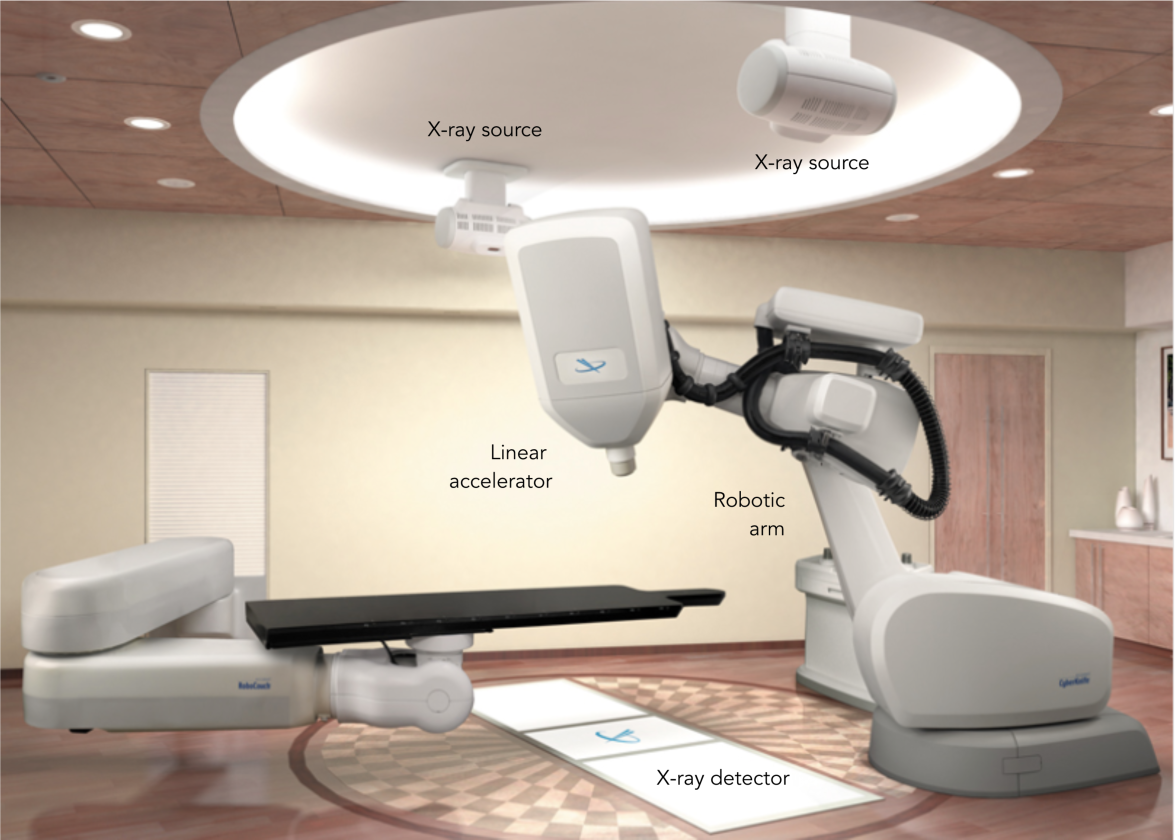
CyberKnife system.

The radiation treatment was performed in an outpatient setting without anesthesia. Mild sedation was allowed, per patient request. During the procedure, the ICD was temporarily programmed to “monitor only” to prevent inappropriate shocks; pacing settings were left unchanged. At the end of the procedure, the fiducial temporary pacing lead was removed by the electrophysiologist and the ICD interrogated to ensure proper function and tachycardia therapies programmed back to their original settings.

For each treatment plan, the following metrics were taken into consideration:^15^

- dosimetry:
  - minimum dose, mean dose, and maximum dose
  - prescription isodose: % of the maximum dose chosen to be delivered at the periphery of the PTV
- quality of target coverage
  - percentage target covered (PTC): % of the PTV covered by the chosen prescription dose)
  - conformity index (CI): prescription isodose volume (PIV - or, volume that receives the chosen prescription dose)/PTV; and new conformity index (nCI): PIV*PTV/(volume of the PTV covered by PIV)^2^
  - homogeneity index (HI): maximum dose/prescription dose

In simpler terms, CI and nCI (the latter better suited for non-symmetrical PTVs) describe the degree to which the volume that receives the chosen prescription dose conforms to the shape and size of the PTV, thus expressing the amount of healthy tissue dose, whereas HI measures the uniformity of dose within the PTV (Figure 3).

**Figure 3.**
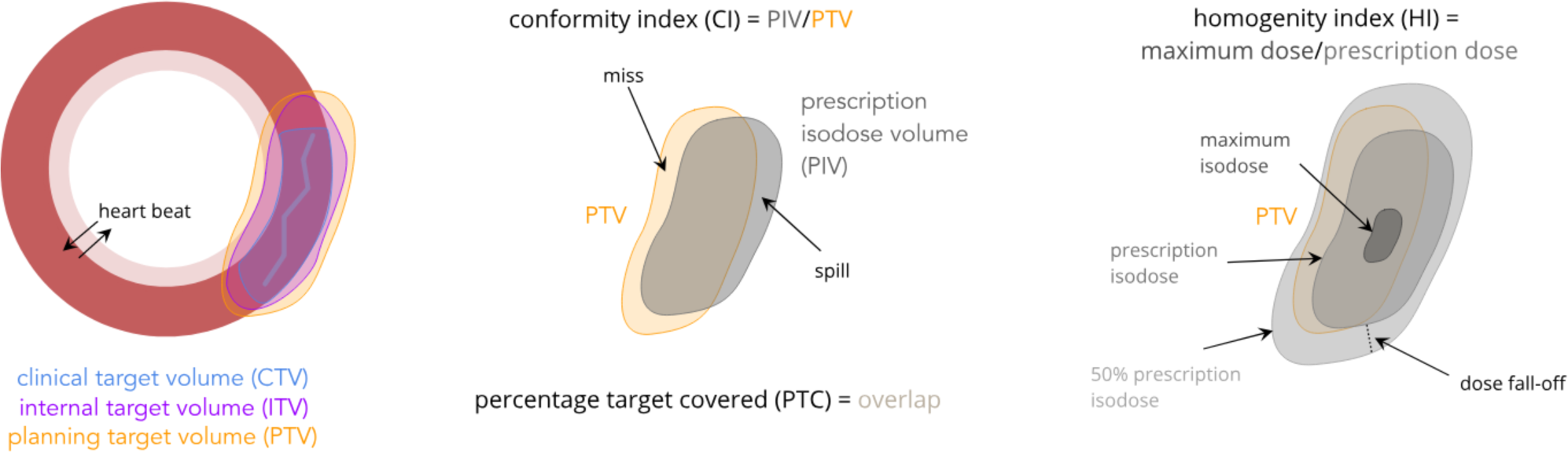
Illustration of basic radiotherapy terms.

- delivery efficiency
  - non-zero beans: photon beans used during treatment to deliver the prescribed dose
  - monitor units (MU): amount of charge recorded in the ionization chamber during treatment
  - treatment time

### Follow-up

After radioablation, patients underwent clinical follow-up scheduled at 2 weeks, 1 month, 2 months, 3 months, 6 months, 9 months and 1 year, at which times they were evaluated for ventricular arrhythmias control (efficacy) and serious clinical events (safety). Arrhythmia control was evaluated by looking at freedom from ventricular arrhythmias and number of AADs required at follow-up.

Ventricular arrhythmia recurrence was defined as presence of VT/ventricular fibrillation receiving appropriate ICD treatment (shock and anti-tachycardia pacing) or documented by ECG/device intracardiac recordings for VTs below the programmed device cut-off. ICD follow-up was performed via remote monitoring in 3/5 patients, while interrogations performed at the time of the clinical follow-up visits and hospitalizations were used in the remaining 2 patients. An electrophysiologist adjudicated all VT episodes (ICD interrogations/ECG). Of note, ICD programming was kept consistent before and after the STAR procedure. AADs prescribed at follow-up were either continued at the same dose, continued at a reduced dose, or discontinued per the treating electrophysiologist.

To assess safety, patients were monitored clinically and with serial imaging for serious thoracic adverse events, with particular attention for Radiation Therapy Oncology Group/European Organization for Research and Treatment of Cancer (RTOG/EORTC) Late Morbidity Scoring Scheme grade ≥ 3 radiation-related toxicity of OAR (i.e. signs and symptoms secondary to inflammation, fibrosis or necrosis in the aforementioned organs). ^16^ Serial imaging consisted of transthoracic echocardiography (TTE - scheduled at 1 month, 6 months, 9 months, and 12 months), chest X-ray (CXR - scheduled at 1 month, 2 months, 6 months.

### Statistical Analysis

Statistical analysis was performed with GraphPad Prism 6.0 for Mac (GraphPad Software, La Jolla, CA, USA). Continuous variables are presented as mean ± standard deviation (SD) or median and range, as appropriate; categorical variables are expressed as percentages.

## Results

Between January and June 2018, 6 patients were screened and enrolled in the study in one center. One patient was subsequently excluded due to pre-existing chronic renal failure, with inability to undergo serial CT imaging with contrast at follow-up. Five patients underwent STAR, their demographic and clinical characteristics are outlined in Table 1. An enrollment exception was made for two patients aged < 60 years old, one of which experienced recurrent sustained symptomatic VT episodes below the ICD therapy cut-off. All patients were males, with a mean age of 63±13 years. The underlying cardiomyopathy was ischemic in 4 (80%), with a mean LVEF of 34±15 %. All patient had scar-related VT refractory to RFCA (scar homogenization), taking an average of 2 (1-2) AADs at the time of enrollment (all having failed amiodarone therapy). Of note, 3 out of 4 patients with ischemic cardiomyopathy underwent prior sternotomy, which prevented epicardial RFCA.

**Table 1.** Clinical characteristics

|  | Gender | Age (years) | Cardiomyopathy | LVEF (%) | NYHA Class | Sternotomy | RFCA | AAD* |
| --- | --- | --- | --- | --- | --- | --- | --- | --- |
| 1 | M | 70 | ischemic | 20 | II | Yes | 1 | 2 |
| 2 | M | 45 | ischemic | 25 | II | Yes | 1 | 2 |
| 3 | M | 55 | ischemic | 55 | I | No | 1° | 1 |
| 4 | M | 67 | ischemic | 25 | II | Yes | 2 | 2 |
| 5 | M | 76 | non-ischemic | 45 | II | Yes | 2 | 1 |

AAD, antiarrhythmic drugs; LVEF, left ventricular ejection fraction; M, male; NYHA, New York Heart Association; RFCA, radiofrequency catheter ablation
\*Vaughan Williams Class I and III; of note, all patients failed amiodarone
°with epicardial access, but no evidence of scar on voltage mapping

Characteristics of the procedure, including the location of the targeted scar and associated delivery efficiency values are in Table 2, whereas detailed data on dosimetry and quality of target coverage are in Table 3. The mean volume of the PTV was 143 ± 50 mL, treated using a mean of 185 ± 48 non-zero beans and 24942 ± 2773 MU over 82 ± 11 minutes. The mean radiation dose delivered to the PTV ranged between 2450 and 2828 cGy, with a mean of 2686 cGy. This dose was prescribed to a 74-80 (mean 77) % isodose line, covering 82 ± 17 % of the PTV. The maximum and minimum dose ranged between 3125 and 2278 (mean 3242) cGy, and 719 and 2076 (mean 1474) cGy, respectively. Mean CI, nCI, and HI were 1.17, 1.50, and 1.30, respectively. STAR was uneventful, and all patients were discharged home the same day without acute procedure-related complications, including normal ICD generator and lead function.

**Table 2.** Procedural characteristics

|  | Location of targeted scar | PTV (mL) | Non-zero beams | Total MU | Time (min) |
| --- | --- | --- | --- | --- | --- |
| 1 | Basal and mid-inferior, apical inferior | 96 | 224 | 26141 | 89 |
| 2 | Anterior mid-apical, septal mid-apical | 184 | 195 | 22388 | 87 |
| 3 | Anterior mid* | 173 | 162 | 24404 | 77 |
| 4 | Anterior mid-apical, septal mid-apical | 180 | 228 | 29107 | 92 |
| 5 | Septal, basal-mid* | 80 | 114 | 22669 | 66 |

MU, monitor units, PTV, planning target volume
\*intramyocardial

**Table 3.** Dosimetry and quality of target coverage

|  | Dose (cGy) |  |  | Prescription isodose (%) | PTC (%) | CI | nCI | HI |
| --- | --- | --- | --- | --- | --- | --- | --- | --- |
|  | Minimum | Mean | Maximum |  |  |  |  |  |
| 1 | 1055 | 2563 | 3125 | 80 | 66* | 1.27 | 1.92 | 1.25 |
| 2 | 1986 | 2828 | 3378 | 74 | 94 | 1.11 | 1.23 | 1.35 |
| 3 | 2076 | 2765 | 3333 | 75 | 91 | 1.12 | 1.22 | 1.33 |
| 4 | 1535 | 2823 | 3247 | 77 | 95 | 1.18 | 1.24 | 1.30 |
| 5 | 719 | 2450 | 3125 | 80 | 61° | 1.15 | 1.89 | 1.25 |

CI, conformity index; HI, heterogeneity index; nCI, new conformity index; PTC, percentage target covered
\*dose reduced to limit radiation exposure to the stomach
°dose reduced to limit radiation exposure to the proximal conduction system

Clinical follow-up ranged from 10 to 14 months, with a mean of 12 ± 2 months. Efficacy results are outlined in Figure 4 and Table 4. Four patients had a marked reduction in ventricular arrhythmia burden during the first 6 months following the procedure, as evidenced by the reduction of VT episodes (both requiring ICD intervention and monitored) and an overall trend in reduction/discontinuation of the baseline AAD. After doing well, thus reducing the AAD dose, patient #5 experienced clinically significant VT recurrence at 3 months, requiring repeated ICD interventions (both shocks and ATP). This patient had a septal scar, which was sub-optimally covered by irradiation (61% PTC) given the vicinity of the atrio-ventricular node. By the end of follow-up, all patients had clinically significant VT recurrence requiring reinstatement of previously tapered down/stopped AAD. Moreover, 3 patients underwent repeat RFCA. Of note, in all patients, voltage mapping has shown areas of surviving myocardium well within the dense scar that was part of the putative PTV. In patient #2, where the electroanatomical voltage maps performed before and after STAR were available for comparison, we also observed an increase of the scar area beyond the boundaries of prior ablation (Figure 5).

**Table 4.**
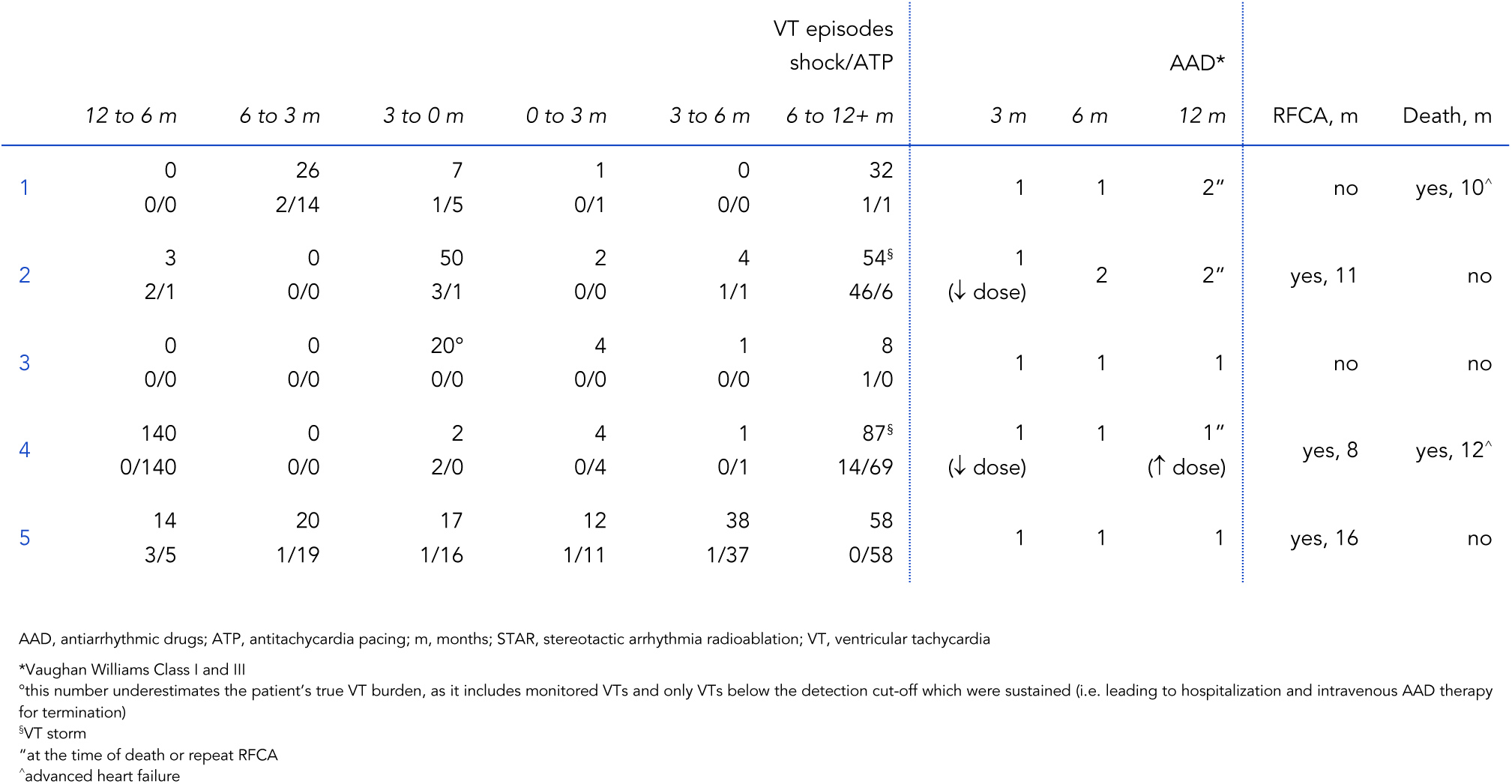
Follow-up

**Figure 4.**
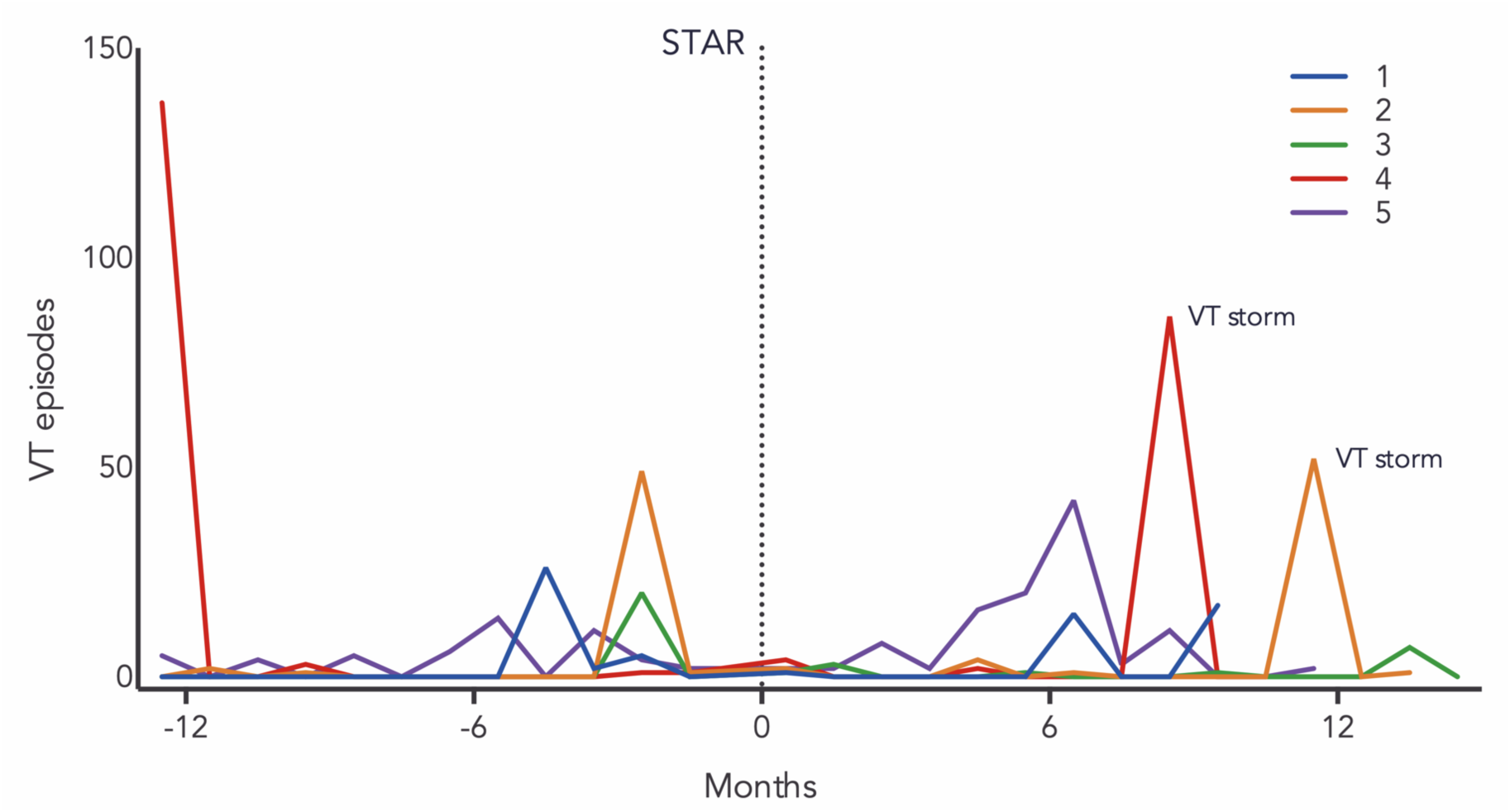
Ventricular arrhythmia burden over time in the patient population, Number of VT episodes per patient in the 12 months prior to and after STAR (dotted line). STAR, stereotactic arrhythmia radioablation; VT, ventricular tachycardia

**Figure 5.**
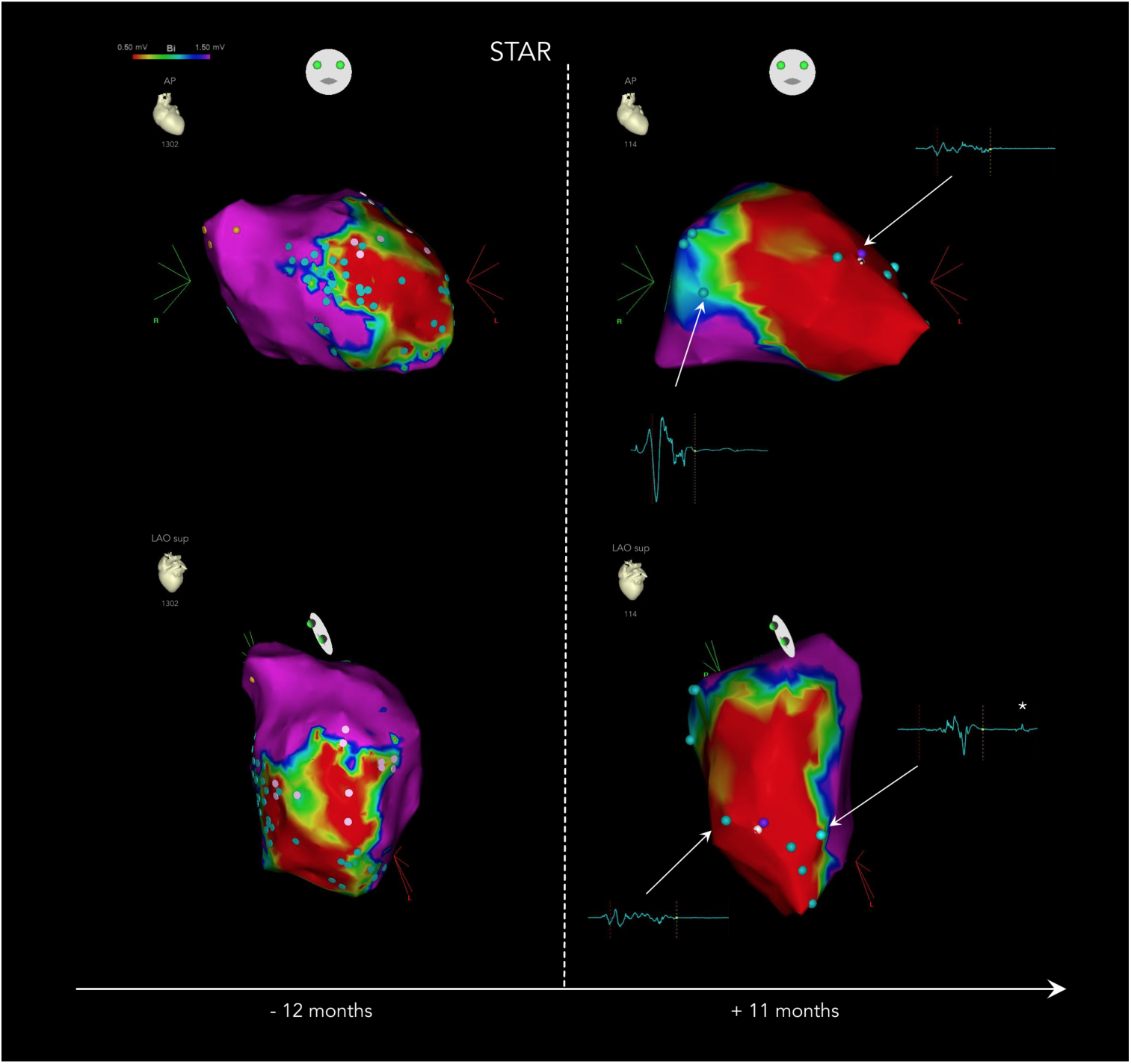
Voltage mapping before and after STAR. Electroanatomical bipolar voltage maps of patient #2 before and after STAR. In the latter, the area of low voltage extends beyond previous borders, with fragmented, late (*) and low voltage potentials within the dense scar. The before-map was obtained using a multipolar catheter with automatic point acquisition, the after-map was obtained in a point by point fashion using a remote magnetic navigation ablation catheter, with manual acquisition and review of each mapping point. AP, anteroposterior; Bi, bipolar; LAO sup, left anterior oblique, superior view; STAR, stereotactic arrhythmia radioablation

There was no clinical or imaging evidence of radiation necrosis or other radiation-induced complications in the OAR. Of note, due to non-compliance, imaging follow-up was not complete in patient #2 (6- and 9-months scheduled exams; 12-months TTE), patient #3 (12-months chest CT scan), and patient #5 (9-months CXR). These patients underwent scheduled imaging follow-up at 18 months (consisting of CXR, TTE and chest CT scan), which confirmed absence of radiation-induced complications in the OAR. LVEF remained stable throughout follow-up in 3 patients, with a transient reduction noted in patient #3 and #5. In the former, the 12-month TTE showed a LVEF of 30% (baseline 55%): this exam was performed at the time of his VT recurrence (failed ATP, with degeneration to VF, requiring 3 shocks for termination), and LVEF improved back to baseline by the 18-month TTE follow-up. In the latter, the 9-months TTE showed a smaller reduction of LVEF (35%, baseline 45%), which returned to baseline by the time of the 12-month TTE follow-up. Two patients (patient #1 and #4) died at 10 and 12 months after the procedure due complications related to their advanced heart failure; no autopsy was performed.

## Discussion

This prospective pilot study demonstrates that, despite good initial results, STAR with CyberKnife did not result in effective ventricular arrhythmia control in the long term in a selected, high-risk population of patients with scar-related VT.

### Stereotactic arrhythmia radioablation

Scar homogenization with RFCA is widely used to treat VT, however there are some anatomical constraints (i.e. deep myocardial origin, unattainable epicardial access) that are known to hinder its efficacy. In addition, RFCA is associated with a significant number of possible procedure-related complications (such as vascular and myocardial injury, embolism, radiation exposure, myocardial ischemia and heart failure), with further limit its application to fragile patients. Thus, patients continuing to have VT who fail or who are unsuitable for RFCA and AADs have limited options. Among the alternative approaches used in clinical practice to overcome these limitations, STAR has recently emerged with promising results.^3–10^ STAR is the application of stereotactic radiosurgery in cardiac arrhythmias. Stereotactic radiosurgery is a non-invasive technology that has been successfully used to treat a variety of solid tumors in the head and neck, lung, abdomen, pelvis and spine, with good local control rates and minimal side effects.^2^ Stereotactic radiosurgery consists of one fraction of very high-dose radiotherapy delivered to the target with submillimeter. The mechanism of injury is postulated to be a combination of vascular injury (leading to tissue hypoxia and necrosis) and apoptotic cell death (resulting in fibrosis and scar formation), which occur over days to months.^1^ Since this is not a cell-specific effect, it is important to concentrate radiation exposure to the target, with a rapid dose falloff to minimize toxicity to surrounding tissue. Histologic and physiologic data have shown that stereotactic radiosurgery is able to create a lesion comparable to the one obtained by RFCA using a dose of 25 to 35 Gy delivered in a single fraction.^17–19^ STAR is a promising technique, because its eliminates risks of RFCA and its associated hospitalization. Early reported outcomes show no procedural-related complications and a decreased arrhythmia burden at follow-up, but additional clinical experience is needed to further define its efficacy and safety.

### Efficacy

In this pilot study, STAR performed with CyberKnife, resulted in an initial decrease in ventricular arrhythmia burden in all patients, but significant recurrence was ineluctable by the end of the 1-year follow-up. It is not easy to explain these disappointing results, but we can hypothesize biological, technical and clinical reasons, which could have – alone or in synergy – influenced the outcomes.

#### Biological considerations

While 25 Gy is the dose used in current clinical studies, pre-clinical dose studies have shown that doses > 30 Gy to the PTV are required to achieve consistent scar formation at 6 months.^20,21^ Underdosing might have led to an initial beneficial effect due to acute (vasogenic and cytokine-release related) edema, but recurrence in the long term due to lack of uniform myocardial cell death.^22^ However, given the current technical limitations (see below), delivering higher target doses while trying to spare the surrounding organs might be not safe, and longer term safety outcomes are necessary before dose escalation. It is also possible that a dose of 25 Gy is adequate, but longer times (> 12 months) are required for transmural scar formation in a tissue mostly composed of terminally differentiated cells, yielding to a higher arrhythmic risk in the mid to long term, during which increased AAD coverage might be necessary. While early (< 3 months) histology examination on human hearts after STAR using 25 Gy has shown no to mild acute inflammatory changes, longer term data are missing.^5,10^ Given these uncertainties, we believe that formal assessment of lesion formation (i.e. serial EAM, biopsy, MRI/CT) on the irradiated myocardial tissue should be incorporated in future human studies to better understand the dynamics of the biological effects of radioablation and more precisely determine the appropriate dosing protocols to optimize safety and efficacy.

#### Technical considerations

Suboptimal radiation coverage is possible, as the myocardial scar is a challenging target for stereotactic radioablation: i) it is not accurately defined by current imaging techniques, ii) it is asymmetrical, and ii) it has complex motion, all while being surrounded by various radio-sensitive organs.

As for the first point, we treated the myocardial scar as defined by areas of myocardial thinning on contrast enhanced first-pass CT correlating with low voltage areas on electroanatomical mapping along with the clinical VT morphology on the 12-lead ECG. Each technique has its limitations, such as inability to detect the border zone, presence of multiple non-scar related factors affecting local voltage, or localization of exit vs critical isthmus sites.^23,24^ While we chose this approach to integrate anatomical and functional information, it might not accurately define the true anatomical extent of the clinically relevant arrhythmogenic substrate to be translated into the radiosurgical target.

Secondly, while contouring was facilitated by CardioPlan, which provided 3-dimensional images and volumes to work with, there are still some challenges associated with the asymmetrical nature of the scar. That is, it is harder to provide uniform irradiation with minimal spillage of significant doses to the surrounding healthy OAR, as evidenced by the non-ideal (> 1) HI and nCI of our treatment plans.^15^ Indeed, in the 2 patients who underwent repeat RFCA, there was evidence of incomplete ablation within the dense scar, with fragmented and delayed potentials noted in tissue well within the irradiated volume.

For the last technical notion, movement of the target results in either the target receiving less than the prescribed dose or the surrounding OAR receiving an additional, unnecessary dose. To avoid the latter, we have followed strict dose limit guidelines, which de facto led to inadequate radiation dosing in 2/5 patients, in whom only about 60% of target was irradiated with 25 Gy. As for the former, the myocardial scar lies in an organ that is constantly moving, itself and within the chest. While there are several techniques to properly follow radiosurgical targets that move along the respiratory cycle, little is known as to how to properly manage cardiac motion. In the current study, we used Synchrony to track the scar in real time: acquiring serial orthogonal X-rays of the fiducial lead located near the target coupled with chest wall motion determined by an optical tracking system, the beam of irradiation moved to coincide with scar throughout the respiratory cycle. Indeed, compared to previous reports, we did not we did not observe inflammatory changes in the surrounding lung parenchyma, possibly signifying a more targeted radioablation with CyberKnife vs other radiotherapy delivery systems.^5^ However, it is possible that by improving the safety profile, arrhythmic outcomes were affected due to more intramyocardial delivery of subtherapeutic doses. As for cardiac motion, we did not have any good real-time indicator to track myocardial movement (i.e. contractility), which was accounted for just by adding a 3 mm margin to the CTV, the static anatomical target that includes the transmural myocardium encompassing the scar. Indeed, previous four-dimensional imaging studies have shown that both the left atrial and ventricular myocardium display significant volumetric, positional and morphological variations throughout the cardiac cycle.^25–27^ Therefore, the lack a direct, real-time, patient-specific internal margin might have led to inadequate/non-uniform irradiation, despite good planned dosimetry and target coverage parameters. This effect is expected to be more pronounced in the periphery of the irradiated volume (penumbra), with an incomplete, non-transmural ablation maturing over time. To support this, in the 2 patients with repeat RFCA, there was also evidence of extension of the scar beyond what was seen in the RFCA preceding radiotherapy. The surviving myocardial tissue as well as the newly maturated border zone are highly arrhythmogenic, leading to VT storm after a period of adequate ventricular arrhythmia control.

#### Clinical considerations

On a clinical standpoint, this is a cohort of patients with advanced heart failure: indeed, 2 patients died of worsening heart failure in the course of follow-up. This is a high-risk population, in which the arrhythmogenic myocardial substrate is large and has already proven to be refractory to available treatments addressing the etiology (anti-ischemic drugs or interventions in the case of ischemic cardiomyopathy), progression (anti-remodeling drugs) and consequences (AADs and RFCA with scar homogenization) of the underlying disease.^28^ Scar progression is expected in this population, and can explain the formation of new arrhythmogenic substrate, which can lead to new ventricular arrhythmias, despite good early outcomes.^29^

### Safety

Finally, we have not observed any acute or early radiation related complications, confirming previous reports using the same dosing protocol (i.e. 25 Gy in a single fraction). Peri-procedural risks are typically extremely low due to the non-invasive nature of treatment. All patients were treated in an outpatient setting, with no complaints or complications. More specifically, we have not observed any ICD generator or lead malfunction, which confirm previous reports of safe irradiation of cardiac hardware up to relatively high energies (> 10 MV).^30^ As for late radiation-necrosis complications, by applying the RGOT validated dose limits, we have not observed any clinical or imaging evidence of significant injury to the OAR up to 1 year of follow-up. However, since late radiation injury risk to the OARs is not only dose dependent, but also reflects the tissue-specific vulnerability, longer surveillance is necessary to confirm this favorable safety profile. As for the transient reduction of LVEF noted in two patients, it can be explained by stunning secondary to the concomitant VT recurrence and ICD shock and mild variation within the natural history/interobserver agreement boundaries. As other patients in our and other reported series had stable LVEF throughout their follow-up, a transient reduction due to radiation injury is less likely, but still possible.^8,9^ More data are necessary to exclude this potential time-dependent adverse effect.

It is also important to note that placement of a temporary lead is an added, albeit small, risk to the procedure. The temporary lead is required for respiratory tracking of the PTV with Synchrony, which requires internal markers for accurate dynamic tracking.^14^ In prior studies, we’ve found that the ICD lead tip is not accurately tracked by Synchrony due to artifacts created by the near right ventricular coil.^31^ On the other hand, the temporary lead is a simple active fixation lead which creates minimal imaging artifacts, and as such is well tracked by Synchrony. The right ventricular septal area is chosen to avoid tracking interference from the patient’s existing ICD lead and to be as close as possible to the target. Of note, three of our patients did have an existing cardiac resynchronization therapy left ventricular lead: dedicated planning study are needed to validate their use for Synchrony tracking, which would remove the need of a temporary lead.

## Conclusion

Despite good initial results, STAR did not result in effective ventricular arrhythmia control in the long term in a selected, high-risk population of patients with scar-related VT. The safety profile of STAR was confirmed to be favorable, with no radiation-related complications observed during the first year of follow-up. Further studies are needed to explain these disappointing results.

## Data Availability

The authors declare that all data supporting the findings of this study are available within the pape

## References

1. Song CW, Kim M-S, Cho LC, Dusenbery K, Sperduto PW. Radiobiological basis of SBRT and SRS. Int J Clin Oncol. 2014;19(4):570–578. doi:10.1007/s10147-014-0717-z.

2. Haridass A. Developments in Stereotactic Body Radiotherapy. Cancers. 2018;10(12):497. doi:10.3390/cancers10120497.

3. Cvek J, Neuwirth R, Knybel L, Molenda L, Otahal B, Pindor J, Murárová M, Kodaj M, Fiala M, Branny M, Feltl D. Cardiac Radiosurgery for Malignant Ventricular Tachycardia. Cureus. 2014;6(7):e190. doi:10.7759/cureus.190.

4. Loo BW, Soltys SG, Wang L, Lo A, Fahimian BP, Iagaru A, Norton L, Shan X, Gardner E, Fogarty T, Maguire P, Al-Ahmad A, Zei P. Stereotactic Ablative Radiotherapy for the Treatment of Refractory Cardiac Ventricular Arrhythmia. Circ Arrhythm Electrophysiol. 2015;8(3):748–750. doi:10.1161/CIRCEP.115.002765.

5. Cuculich PS, Schill MR, Kashani R, Mutic S, Lang A, Cooper D, Faddis M, Gleva M, Noheria A, Smith TW, Hallahan D, Rudy Y, Robinson CG. Noninvasive Cardiac Radiation for Ablation of Ventricular Tachycardia. N Engl J Med. 2017;377(24):2325–2336. doi:10.1056/NEJMoa1613773.

6. Jumeau R, Ozsahin M, Schwitter J, Vallet V, Duclos F, Zeverino M, Moeckli R, Pruvot E, Bourhis J. Rescue procedure for an electrical storm using robotic non-invasive cardiac radio-ablation. Radiother Oncol. 2018;128(2):189–191. doi:10.1016/j.radonc.2018.04.025.

7. Scholz EP, Seidensaal K, Naumann P, André F, Katus HA, Debus J. Risen from the dead: Cardiac stereotactic ablative radiotherapy as last rescue in a patient with refractory ventricular fibrillation storm. Hear Case Rep. 2019;5(6):329–332. doi:10.1016/j.hrcr.2019.03.004.

8. Robinson CG, Samson PP, Moore KMS, Hugo GD, Knutson N, Mutic S, Goddu SM, Lang A, Cooper DH, Faddis M, Noheria A, Smith TW, Woodard PK, Gropler RJ, Hallahan DE, Rudy Y, Cuculich PS. Phase I/II Trial of Electrophysiology-Guided Noninvasive Cardiac Radioablation for Ventricular Tachycardia. Circulation. 2019;139(3):313–321. doi:10.1161/CIRCULATIONAHA.118.038261.

9. Neuwirth R, Cvek J, Knybel L, Jiravsky O, Molenda L, Kodaj M, Fiala M, Peichl P, Feltl D, Januška J, Hecko J, Kautzner J. Stereotactic radiosurgery for ablation of ventricular tachycardia. EP Eur. 2019;21(7):1088–1095. doi: https://doi.org/10.1093/europace/euz133.

10. Lloyd MS, Wight J, Schneider F, Hoskins M, Attia T, Escott C, Lerakis S, Higgins KA. Clinical experience of stereotactic body radiation for refractory ventricular tachycardia in advanced heart failure patients. Heart Rhythm. October 2019. doi:10.1016/j.hrthm.2019.09.028.

11. Benedict SH, Yenice KM, Followill D, Galvin JM, Hinson W, Kavanagh B, Keall P, Lovelock M, Meeks S, Papiez L, Purdie T, Sadagopan R, Schell MC, Salter B, Schlesinger DJ, Shiu AS, Solberg T, Song DY, Stieber V, Timmerman R, Tomé WA, Verellen D, Wang L, Yin F-F. Stereotactic body radiation therapy: The report of AAPM Task Group 101: Stereotactic body radiation therapy: The report of TG101. Med Phys. 2010;37(8):4078–4101. doi:10.1118/1.3438081.

12. Coste-Manière È, Olender D, Kilby W, Schulz RA. Robotic whole body stereotactic radiosurgery: clinical advantages of the Cyberknife® integrated system. Int J Med Robot. 2005;1(2):28–39. doi:10.1002/rcs.39.

13. Ho AK, Fu D, Cotrutz C, Hancock SL, Chang SD, Gibbs IC, Maurer CR, Adler JR. A Study of the Accuracy of CyberKnife Spinal Radiosurgery Using Skeletal Structure Tracking. Oper Neurosurg. 2007;60:147–156. doi:10.1227/01.NEU.0000249248.55923.EC.

14. Ozhasoglu C, Saw CB, Chen H, Burton S, Komanduri K, Yue NJ, Huq SM, Heron DE. Synchrony – Cyberknife Respiratory Compensation Technology. Med Dosim. 2008;33(2):117–123. doi:10.1016/j.meddos.2008.02.004.

15. Torrens M, Chung C, Chung H-T, Hanssens P, Jaffray D, Kemeny A, Larson D, Levivier M, Lindquist C, Lippitz B, Novotny J, Paddick I, Prasad D, Yu CP. Standardization of terminology in stereotactic radiosurgery: Report from the Standardization Committee of the International Leksell Gamma Knife Society. J Neurosurg. 2014;121(Suppl_2):2–15. doi:10.3171/2014.7.GKS141199.

16. Cox JD, Stetz J, Pajak TF. Toxicity criteria of the Radiation Therapy Oncology Group (RTOG) and the European organization for research and treatment of cancer (EORTC). Int J Radiat Oncol. 1995;31(5):1341–1346. doi:10.1016/0360-3016(95)00060-C.

17. Sharma A, Wong D, Weidlich G, Fogarty T, Jack A, Sumanaweera T, Maguire P. Noninvasive stereotactic radiosurgery (CyberHeart) for creation of ablation lesions in the atrium. Heart Rhythm. 2010;7(6):802–810. doi:10.1016/j.hrthm.2010.02.010.

18. Lehmann HI, Deisher AJ, Takami M, Kruse JJ, Song L, Anderson SE, Cusma JT, Parker KD, Johnson SB, Asirvatham SJ, Miller RC, Herman MG, Packer DL. External Arrhythmia Ablation Using Photon Beams: Ablation of the Atrioventricular Junction in an Intact Animal Model. Circ Arrhythm Electrophysiol. 2017;10(4):e004304. doi:10.1161/CIRCEP.116.004304.

19. Zei PC, Wong D, Gardner E, Fogarty T, Maguire P. Safety and efficacy of stereotactic radioablation targeting pulmonary vein tissues in an experimental model. Heart Rhythm. 2018;15(9):1420–1427. doi:10.1016/j.hrthm.2018.04.015.

20. Blanck O, Bode F, Gebhard M, Hunold P, Brandt S, Bruder R, Grossherr M, Vonthein R, Rades D, Dunst J. Dose-Escalation Study for Cardiac Radiosurgery in a Porcine Model. Int J Radiat Oncol. 2014;89(3):590–598. doi:10.1016/j.ijrobp.2014.02.036.

21. Lehmann HI, Graeff C, Simoniello P, Constantinescu A, Takami M, Lugenbiel P, Richter D, Eichhorn A, Prall M, Kaderka R, Fiedler F, Helmbrecht S, Fournier C, Erbeldinger N, Rahm A-K, Rivinius R, Thomas D, Katus HA, Johnson SB, Parker KD, Debus J, Asirvatham SJ, Bert C, Durante M, Packer DL. Feasibility Study on Cardiac Arrhythmia Ablation Using High-Energy Heavy Ion Beams. Sci Rep. 2016;6(1):38895. doi:10.1038/srep38895.

22. Tapio S. Pathology and biology of radiation-induced cardiac disease. J Radiat Res (Tokyo). 2016;57(5):439–448. doi:10.1093/jrr/rrw064.

23. Komatsu Y, Cochet H, Jadidi A, Sacher F, Shah A, Derval N, Scherr D, Pascale P, Roten L, Denis A, Ramoul K, Miyazaki S, Daly M, Riffaud M, Sermesant M, Relan J, Ayache N, Kim S, Montaudon M, Laurent F, Hocini M, Haïssaguerre M, Jaïs P. Regional Myocardial Wall Thinning at Multidetector Computed Tomography Correlates to Arrhythmogenic Substrate in Postinfarction Ventricular Tachycardia: Assessment of Structural and Electrical Substrate. Circ Arrhythm Electrophysiol. 2013;6(2):342–350. doi:10.1161/CIRCEP.112.000191.

24. Josephson ME, Anter E. Substrate Mapping for Ventricular Tachycardia Assumptions and Misconceptions. JACC Clin Electrophysiol. 2015;1(5):341–352. doi:10.1016/j.jacep.2015.09.001.

25. Ipsen S, Blanck O, Lowther NJ, Liney GP, Rai R, Bode F, Dunst J, Schweikard A, Keall PJ. Towards real-time MRI-guided 3D localization of deforming targets for non-invasive cardiac radiosurgery. Phys Med Biol. 2016;61(22):7848–7863. doi:10.1088/0031-9155/61/22/7848.

26. Tong Y, Yin Y, Lu J, Liu T, Chen J, Cheng P, Gong G. Quantification of heart, pericardium, and left ventricular myocardium movements during the cardiac cycle for thoracic tumor radiotherapy. OncoTargets Ther. 2018;11:547–554. doi:10.2147/OTT.S155680.

27. Hasnain A, Suzuki A, Wang S, Konishi H, Newman L, Parker K, Rettmann ME, Deisher A, Packer D, Hohmann S. Quantitative assessment of cardiac motion using multiphase computed tomography imaging with application to cardiac ablation therapy. In: Webster RJ, Fei B, eds. Medical Imaging 2018: Image-Guided Procedures, Robotic Interventions, and Modeling. Vol 10576. Houston, United States: SPIE; 2018:105762F. doi:10.1117/12.2295438.

28. Cohn JN, Ferrari R, Sharpe N. Cardiac remodeling—concepts and clinical implications: a consensus paper from an international forum on cardiac remodeling. J Am Coll Cardiol. 2000;35(3):569–582. doi:10.1016/S0735-1097(99)00630-0.

29. Berte B, Sacher F, Venlet J, Andreu D, Mahida S, Aldhoon B, De Potter T, Sarkozy A, Tavernier R, Andronache M, Deneke T, Kautzner J, Berruezo A, Cochet H, Zeppenfeld K, Jaïs P. VT Recurrence After Ablation: Incomplete Ablation or Disease Progression? A Multicentric European Study: NICM VT Ablation. J Cardiovasc Electrophysiol. 2016;27(1):80–87. doi:10.1111/jce.12858.

30. Aslian H, Kron T, Longo F, Rad R, Severgnini M. A review and analysis of stereotactic body radiotherapy and radiosurgery of patients with cardiac implantable electronic devices. Australas Phys Eng Sci Med. 2019;42(2):415–425. doi:10.1007/s13246-019-00751-8.

31. Wang L, Fahimian B, Soltys SG, Zei P, Lo A, Gardner EA, Maguire PJ, Loo BW. Stereotactic Arrhythmia Radioablation (STAR) of Ventricular Tachycardia: A Treatment Planning Study. Cureus. 2016;8(7):e694. doi:10.7759/cureus.694.

